# Urogenital Schistosomiasis in Dam-Adjacent Communities, Kano State, Nigeria

**DOI:** 10.64898/2026.08.14.26359244

**Authors:** Mahmud Umar Ali, Sodangi Abdulkareem Luka, Iliya Samuel Ndams, Ezekiel Kogi, Kabir Junaidu, Gloria Dada Chechet, Emmanuel Oluwadare Balogun, Soerge Kelm, Syafinaz Amin-Nordin

## Abstract

**Background:** Urogenital schistosomiasis remains a major neglected tropical disease in rural communities, particularly in reservoir-adjacent settings where sustained human-water contact facilitates persistent transmission. Despite ongoing control efforts, communities located around reservoirs may experience persistent transmission partly due to socioeconomic water exposure and presence of infected intermediate host snails in aquatic habitats. This study assessed the prevalence, intensity, and epidemiological correlates of *Schistosoma haematobium* infection in reservoir-adjacent communities in Kano State, Nigeria to better understand local transmission.

**Methods:** A community-based cross-sectional study was conducted among 447 consenting participants from six communities located around three water reservoirs in Kano State.

Sociodemographic characteristics, water-contact behaviour, sanitation practices, treatment-seeking behaviour, and knowledge related to urogenital schistosomiasis were obtained using interviewer-administered questionnaires. Urine samples collected between 10:00 and 14:00 were examined using microscopy following centrifugation/sedimentation. *Schistosoma haematobium* isolates were characterised by molecular methods, using PCR and Sanger dideoxy sequencing. *Bulinus* snails were collected from the reservoirs by handpicking and scooping methods. Nucleotide sequences were analysed using NCBI BLAST to confirm species identity.

Epidemiological data on *Schistosoma haematobium* infection were analysed using chi-square tests and univariate and multivariate logistic regression analyses to assess associations with explanatory variables.

**Results:** The overall prevalence of *S. haematobium* infection was 34.23% (153/447), indicating moderate endemicity. Molecular study confirmed the occurrence of pure *S. haematobium*.

Community-specific prevalence ranged from 32.35% to 37.84%. Heavy-intensity infection (>50 eggs/10 mL urine) occurred in 17.65% of infected participants. Infection was most common among children and adolescents and was higher in males than females, although the sex difference was not statistically significant. Water-contact behaviour was the strongest correlate of infection: participants reporting water contact had a markedly higher prevalence than those without such exposure (39.12% vs 3.28%; OR = 18.96, 95% CI: 4.56-78.73). Fetching water (aOR = 2.57; 95% CI = 1.53-4.34; p < 0.001) and Swimming (aOR = 2.37; 95% CI = 1.42 - 3.94; p = 0.001) were the specific activities most strongly associated with higher odds of *S. haematobium* infection. Age (aOR = 0.91; 95% CI = 0.87 - 0.95; p < 0.001) significantly reduced the odds of infection. In terms of gender, female participants had significantly lower odds of infection compared with males (aOR = 0.52; 0.29 0.92; p = 0.03). Knowledge of transmission was poor, with only a small proportion of participants correctly identifying contact with contaminated water as the cause of infection. A rare ectopic detection of *S. mansoni* eggs in urine was also observed. Presence of *S. haematobium*-infected *Bulinus* intermediate hosts in the water reservoirs was reported.

**Conclusions:** Urogenital schistosomiasis remains actively transmitted in reservoir-adjacent communities in Kano State, with infection strongly associated with frequent human-water contact and poor disease knowledge. These findings support the need for integrated control strategies combining praziquantel-based preventive chemotherapy, targeted health education, improved water and sanitation, and behaviour-focused interventions to reduce reinfection and interrupt transmission.

**Author summary:** Urogenital schistosomiasis is a neglected tropical disease that continues to affect rural communities in Nigeria, especially those living near water reservoirs where people frequently come into contact with unsafe water. This study investigated how common the infection is and the factors that contribute to its spread in communities around reservoirs in Kano State. We found that about one in three people was infected, indicating that the disease is still actively transmitted. Molecular analysis of *S. haematobium* isolates confirmed species identity. Infection was more common among children and adolescents and was strongly linked to activities such as swimming and fetching water from contaminated sources. Many participants had limited knowledge of how the disease is transmitted, which may contribute to continued exposure and reinfection. The presence of infected snail hosts in the reservoirs further supports ongoing transmission in these communities. These findings highlight the need for improved control measures, including regular treatment with praziquantel, better access to safe water and sanitation, and increased community education to reduce risky water-contact behaviour and interrupt disease transmission.

## Introduction

Human schistosomiasis is a water-borne neglected tropical disease (NTD) caused by the blood flukes of *Schistosoma* spp. According to recent estimates, approximately 779 million people are at risk of schistosomiasis globally, while about 250 million people are infected (Balogun *et al*., 2022; WHO, 2023; Obijiofor *et al*., 2024). The disease disproportionately affects populations living in rural and deprived urban communities where access to clean water and adequate sanitation is limited. In Africa, schistosomiasis is endemic in 78 countries; with an estimated 207 million people infected and about 400 million at risk. Most affected individuals are engaged in agricultural activities, domestic chores, or recreational practices that expose them to cercaria-infested water (Dawaki *et al*., 2015; WHO, 2023).

Human schistosomiasis is mainly caused by five species of the genus *Schistosoma*, namely *S. haematobium, S. mansoni, S. japonicum, S. intercalatum*, and *S. mekongi* (WHO, 2023). Among these, *S. haematobium* is responsible for urogenital schistosomiasis (UgS), which is particularly widespread in sub-Saharan Africa. Urogenital schistosomiasis is one of the most important forms of human schistosomiasis in sub-Saharan Africa.

The global burden of UgS is considerable, with about 100 million people affected, most of whom live in sub-Saharan Africa (Nordin *et al*., 2023). Urogenital schistosomiasis is predominantly distributed in sub-Saharan Africa, with countries such as Nigeria, Tanzania, and Uganda among the most heavily affected. In Africa, more than 30 countries are affected, and the highest numbers of combined *S. haematobium* and *S. mansoni* infections have been reported in Nigeria, Tanzania, Ghana, and Mozambique (Bajiro *et al*., 2016). Infection with *Schistosoma haematobium* has serious economic and social consequences, particularly in rural communities where education is limited, sanitary conditions are poor, and access to healthcare is inadequate (Ali and Ndams, 2013; Dawaki *et al*., 2016).

Despite repeated praziquantel campaigns in Nigeria, transmission of UgS persists in reservoir-dependent rural communities where ecological, behavioural, and malacological determinants intersect. Few studies in northern Nigeria have simultaneously integrated parasitological, molecular, behavioural, and intermediate-host surveillance data to characterise ongoing transmission dynamics. Understanding these interacting determinants is essential for designing context-specific interventions aligned with the WHO 2030 schistosomiasis elimination roadmap.

## Materials and Methods

### Study area and population

The study was conducted in six selected communities located in rural areas around three water reservoirs in Kano State, Nigeria. The communities included Badau (12.1846°N, 8.1256°E) and Daddauda (12.1921°N, 8.1578°E) around Bagwai Dam in Bagwai Local Government Area (LGA); Gamji (11.5981°N, 8.8425°E) and Maimanda (11.6107°N, 8.8415°E) around Kafin Chiri Dam in Garko LGA; and Bangare (12.1227°N, 8.6775°E) and Danmadanhu (12.1007°N, 8.6630°E) around Wasai Dam in Gezawa LGA. These communities are geographically distributed across the state and represent the three senatorial zones of Kano State - Kano North, Kano Central, and Kano South, respectively. The study area lies within the geographical extent of approximately 11°35′55″N - 12°19′37″N and 8°13′52″E - 8°50′20″E (Figure 1).

**Figure 1:**
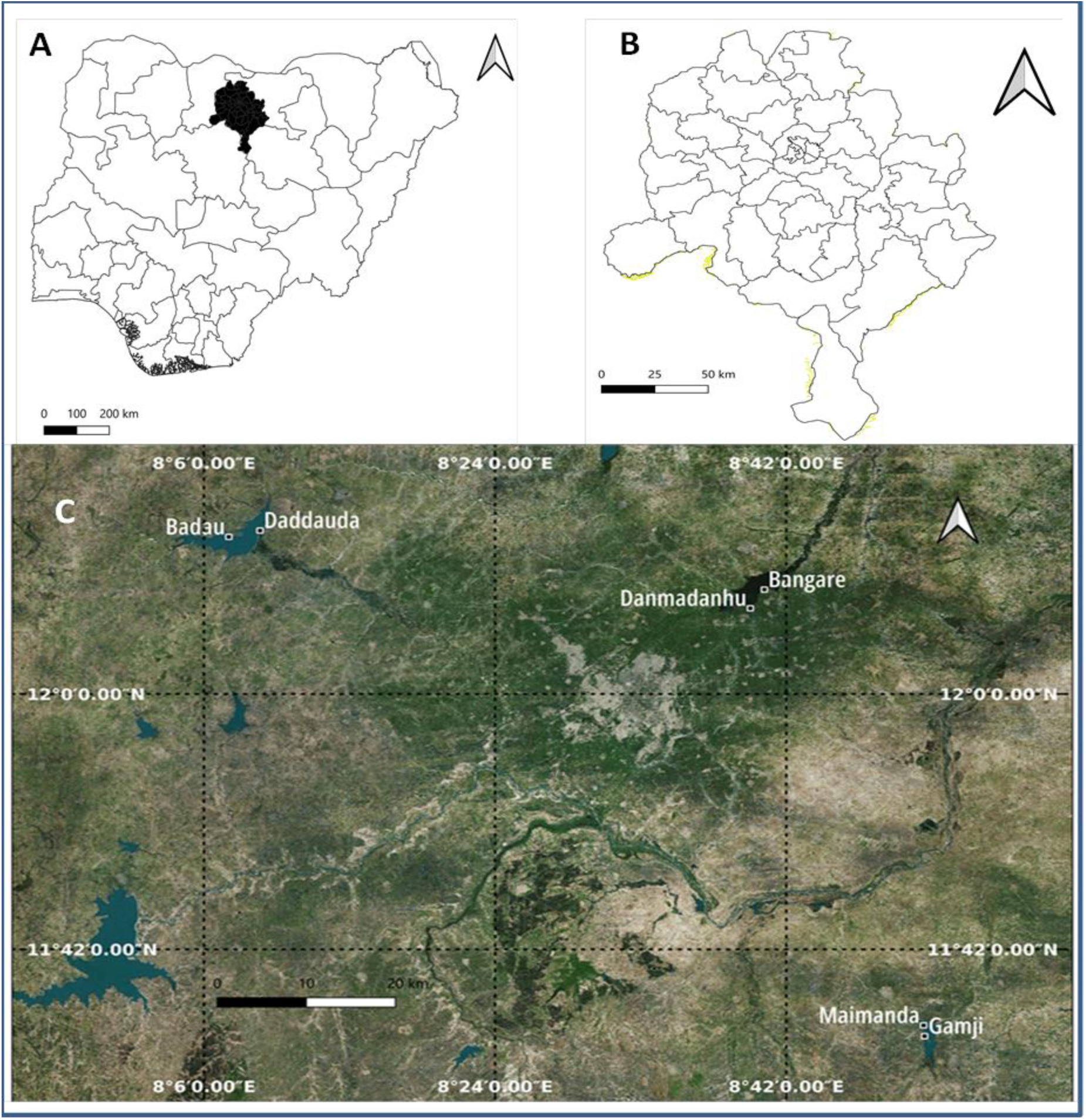
Map of the study area showing: (A) the location of Kano State in Nigeria; (B) Kano State; and (C) the study sites. The map was generated using QGIS software version 3.40.6.

Kano State is the most populous state in northern Nigeria. It lies within the Sudan Savanna vegetation zone, with features of the Sahel Savanna occurring toward the northern part of the state due to increasing desertification. The climate is hot and semi-arid, with distinct wet and dry seasons and approximately four months of rainfall between June and September. The population is predominantly of Hausa tribe and Islamic faith, with a relatively homogeneous cultural background. The study communities are mainly agrarian, relying on farming, particularly irrigation farming, as well as fishing and other water-related livelihood activities. These socioeconomic activities, together with recreational water contact, predispose residents to schistosomiasis transmission.

### Study design

This was a community-based cross-sectional study conducted in six selected communities situated close to water reservoirs that serve multiple socioeconomic, domestic, and recreational functions for the surrounding populations in Kano State. Community selection was based on three criteria: (i) remoteness of the village, defined as being hard-to-reach; (ii) proximity to a water reservoir; and (iii) evidence of human water contact activities (WCA) observed during preliminary visits. For equity and inclusiveness, both male and female individuals of all age groups were considered eligible for participation.

### Research ethics clearance

The research protocol was developed in accordance with the Declaration of Helsinki (World Medical Association, WMA, 2013) and the International Ethical Guidelines for Biomedical Research Involving Human Subjects (CIOMS, 2016), and followed the guidelines of the Health Research Ethics Committees of Ahmadu Bello University, Zaria and the Institutional Review Board (IRB) of Aliko Dangote University of Science and Technology, Wudil. Ethical approval was obtained from the Health Research Ethics Committee of the Kano State Ministry of Health (Reference No. MOH/Off/797/T.I/1869).

Before participant recruitment, extensive community engagement and advocacy were conducted. With the permission of each *Dagaci* or *Mai Unguwa* (traditional community leader), a town crier (*Sankira*) was asked to make public announcements inviting community members to attend town hall meetings held at the residence of each community leader. Imams were also requested to make similar announcements during prayers to encourage community participation.

At these meetings, the purpose, procedures, and potential benefits of the study were explained to the community members. Participants were informed that the study posed minimal or no risk and that they had the right to decline participation or withdraw at any time without any consequences. Confidentiality and privacy of personal information were assured. Individuals who gave informed consent were recruited and provided with clean, labelled sample bottles for urine collection.

### Inclusion and exclusion criteria

All apparently healthy individuals who gave informed consent were recruited into the study. Individuals who were severely ill, recently received praziquantel treatment, or declined consent were excluded.

### Sample size determination

The sample size was determined using Cochran’s formula as described by Enabulele *et al*. (2021). The calculation was based on a reported prevalence of 37% for UgS in Kano State.

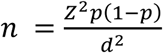

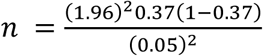

Where:

n= Sample size

Z= 1.96 (at 95% confidence interval)

*P*= 0.37 (Prevalence of UgS in Kano State; Enabulele *et al*. 2021)

d= Margin of error (5% or 0.05)

The calculated minimum sample size was 358. To account for possible participant attrition, at least 10% of the minimum sample size was added, bringing the total target recruitment to 500 participants. This allowed for possible consent withdrawal or incomplete participation without compromising the validity of the study. Ultimately, 447 participants fully complied by providing informed consent, completing the questionnaire, and submitting urine samples for analysis.

### Questionnaire administration

A structured questionnaire was adapted from Ali *et al*. (2021) with minor modifications and pretested before administration. The questionnaire was interviewer-administered through face-to-face interviews and used to collect qualitative data on sociodemographic and epidemiological risk factors for UgS in the study area. Information obtained included age, gender, occupation, level of education, sanitary practices, water-use practices, clinical features, healthcare-seeking behaviour, and knowledge, attitudes, and perceptions (KAP) regarding UgS.

### Collection and analysis of urine samples

Consenting participants were asked to provide clean-catch midstream urine samples between 10:00 a.m. and 2:00 p.m., the period known to coincide with peak egg excretion. Before sample collection, participants were briefly instructed on how to collect uncontaminated urine samples and transport them to the collection point. Each participant received a labelled urine sample bottle. On receipt, the urine samples were preserved in absolute ethanol and transported to the laboratory for microscopy.

Using the concentration method described by Cheesbrough (2009), urine samples were centrifuged at 1500 revolutions per minute for 5 minutes. The supernatant was carefully withdrawn and discarded using a 10 mL syringe fitted with a 21-gauge needle. The remaining sediment was pipetted onto a clean glass slide using a Pasteur pipette, and covered with a coverslip and examined under the ×10 objective of a compound binocular microscope for the characteristic terminal-spined oval eggs of *S. haematobium* (Quan *et al*., 2015). Egg counts were performed systematically to determine infection intensity and recorded as eggs per 10 mL of urine (WHO, 2019).

### Sample preservation

Following centrifugation of urine samples, *S. haematobium* egg pellets were aspirated, transferred into 1.5 mL microcentrifuge tubes, fixed in absolute ethanol, and stored at-20°C until molecular analysis (Tumwebaze *et al*., 2019; El-Kady *et al*., 2020).

### Collection and identification of snail hosts

Freshwater snails were collected from water reservoirs by hand-picking with forceps and by scooping with a long-handled net. During collection, field personnel wore protective gloves and boots to prevent accidental cercarial exposure. Snails were transported to the laboratory in plastic containers containing a small amount of site water and moist vegetation.

Snails were identified morphologically using standard taxonomic keys (Brown and Kristensen, 1993). *Bulinus* species were differentiated by shell spire height and truncation of the columella. *Bulinus globosus* was characterized by a short spire and truncated columella, whereas *B. truncatus* showed a longer spire and no truncation.

### Genomic DNA extraction

Genomic DNA from preserved urine sediments containing *S. haematobium* eggs was extracted using the DNeasy Blood and Tissue Kit (Qiagen, Hilden, Germany) with minor modifications at the Center for Biomolecular Interactions Bremen, University of Bremen, Germany. Briefly, after preserved samples were thawed, 200 µL of urine sediment was centrifuged and washed twice in double-distilled water to remove ethanol. This was then resuspended in phosphate-buffered saline, and lysed with 20 µL proteinase K and buffer AL at 56°C for 1 h. Subsequent purification and elution steps followed the manufacturer’s instructions, and DNA was eluted in 50 µL elution buffer (Zhang *et al*., 2022). DNA concentration and purity were measured using a NanoDrop 1000 spectrophotometer (Thermo Scientific, Dreieich, Germany). DNA concentration was recorded in ng/µL, and purity was assessed using the A260/A280 ratio (Lucena-Aguilar *et al*., 2016).

### PCR amplification

Semi-nested PCR was used to amplify the mitochondrial cytochrome c oxidase subunit 1 (*cox1*) gene of *S. haematobium* (543 bp) using a genus-specific forward primer and an *S. haematobium*-specific reverse primer (F: 5ʹ-TTTTTTGGTCATCCTGAGGTGTAT-3ʹ and R: 5ʹ-TGATAATCAATGACCCTGCAATAA-3ʹ) (Schols *et al*., 2019). PCR reactions were carried out in 20 µL volumes containing 10 µL of 2× DreamTaq Hot Start Green PCR Master Mix (Thermo Scientific), 2 µL each of 1 µM forward and reverse primers, 5 µL of nuclease-free water, and 1 µL DNA template. For *cox1*, thermocycling conditions were: initial denaturation at 95°C for 2 min; 30 cycles of 95°C for 30 s, 58°C for 30 s, and 72°C for 35 s; and final extension at 72°C for 5 min.

### Agarose gel electrophoresis and sequencing

PCR products were resolved on 2% agarose gels stained with 0.3 µL ViSafe Gel Red in 1× TBE buffer (Omran *et al*., 2021). Electrophoresis was performed at 80 V for 50 min, and a 100 bp DNA ladder was used as size standard. Gels were visualized under UV light using transilluminator and subsequently photographed.

Selected amplicons were excised and purified using the GeneJET Gel Extraction Kit (Thermo Scientific, Germany) according to the manufacturer’s instructions. Purified PCR products were quantified, stored at-20°C, and submitted along with genus-specific universal forward primer (5ʹ-TTTTTTGGTCATCCTGAGGTGTAT-3ʹ) for Sanger sequencing at Microsynth Sequencing Laboratories (Göttingen, Germany).

## Data analysis

The epidemiological data generated were coded and entered into Microsoft Excel 2010 and analysed statistically using IBM SPSS version 23 and Epi Info version 7.2.6.0 (CDC).

Prevalence and intensity of *S. haematobium* infections in humans and snail intermediate hosts were calculated using descriptive statistics. Chi-square tests, univariate and multivariate logistic regression analyses were conducted to determine association between sociodemographic risk factors and *S. haematobium* transmission. Statistical significance was set at 95% confidence intervals (CIs) and *P* < 0.05 for all analyses.

In order to confirm species identity and exclude zoonotic or hybrid transmission, two of the positive samples were sequenced by Sanger sequencing technique. Sequence chromatograms were examined using Sequence Scanner version 2 (Applied Biosystems). High-quality sequences were queried against the NCBI nucleotide database using BLASTn, and reference sequences with 99.24%–100% identity were used for species confirmation. The generated nucleotide sequences of *S. haematobium cox1* were subjected to SNP analysis using CodonCode Aligner software (version 12.0.1). Reference sequences for the analysis were downloaded in FASTA format from the NCBI GenBank Database.

## Results

### Sociodemographics of the study participants

The sociodemographic and environmental characteristics of the study participants were presented in Table 1. The study population was predominantly composed of school-aged individuals, with approximately 79.86% aged ≤19 years. The mean age was 15.97 years, with male participants being older on average than females. Males constituted the majority of participants (77.40%).

**Table 1:**
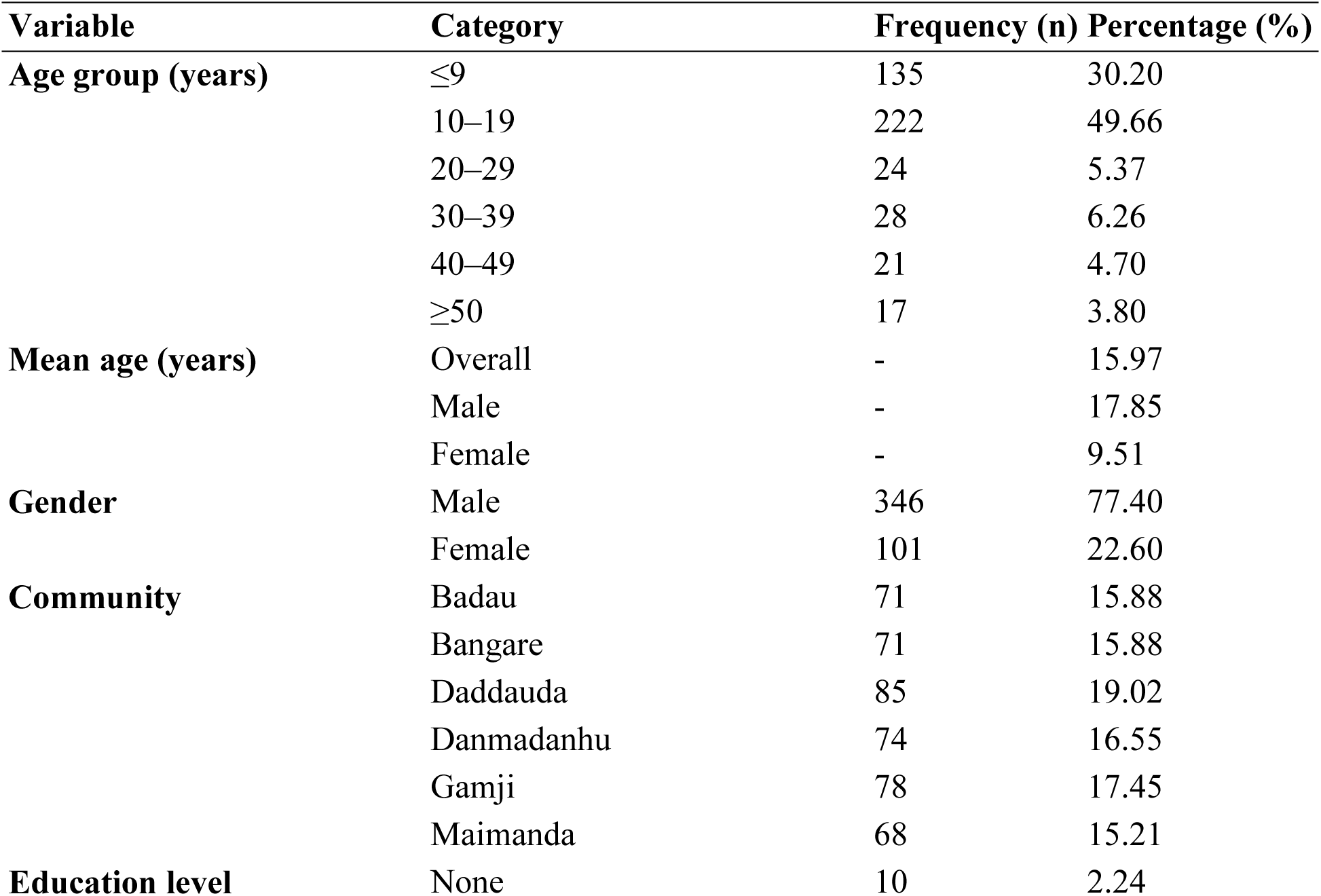
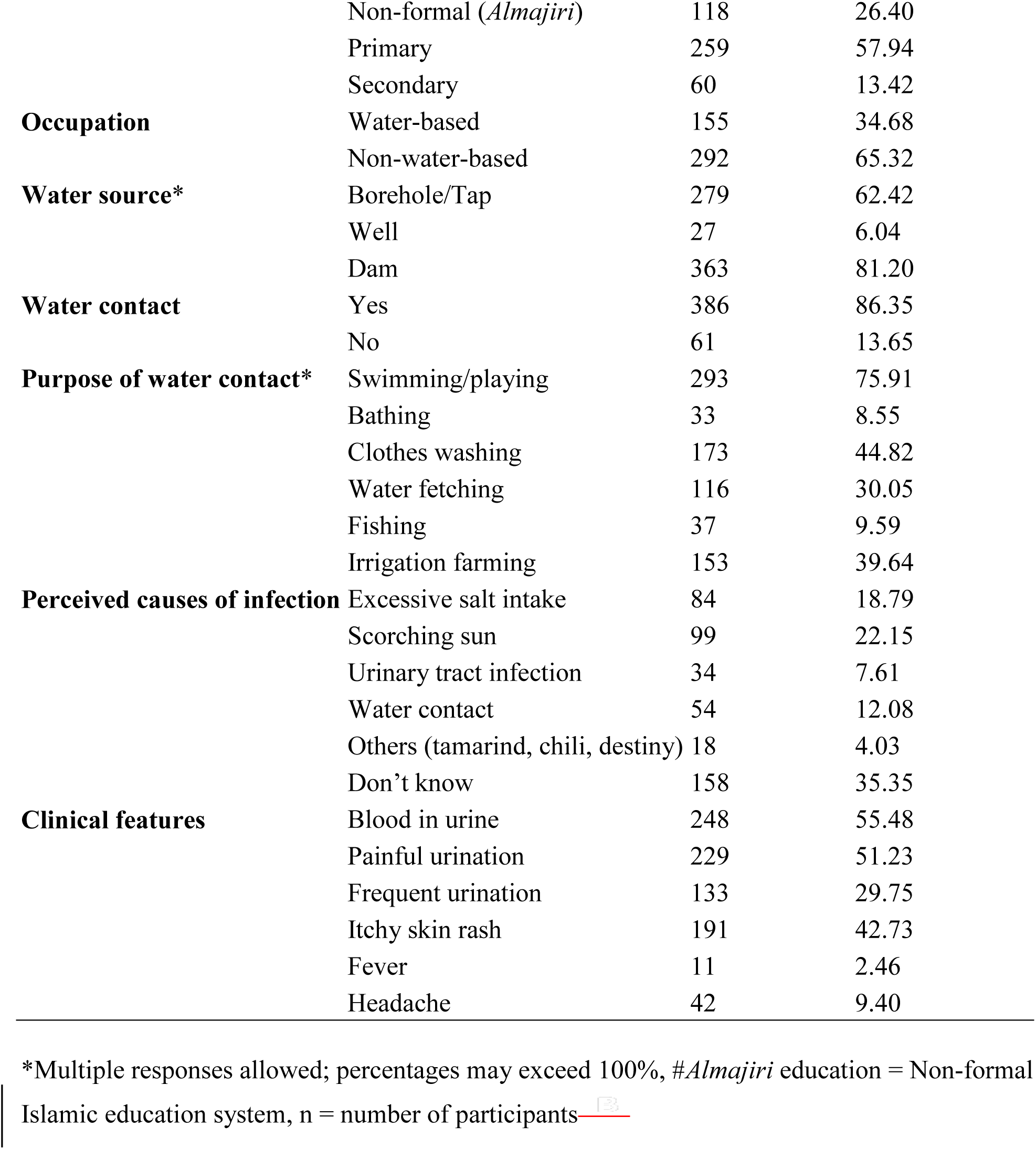
Sociodemographic, environmental, and clinical characteristics of study participants (n = 447)

A high proportion of participants reported exposure to potential transmission sources, with 86.35% having contact with natural water bodies. The primary activities associated with water contact were swimming/playing (75.91%), irrigation farming (39.64%), and clothes washing (44.82%). Frequent occupational and recreational water-contact activities were observed across the study communities, including swimming, fishing, irrigation farming, and domestic utilization of dam water, particularly among school-aged children. These activities may contribute substantially to sustained schistosomiasis transmission within the endemic communities. Most participants relied on potentially unsafe water sources, particularly dams (81.20%), highlighting significant environmental exposure risk. Educational attainment was generally low, with the majority having only primary education (57.94%) or non-formal education (26.40%).

Knowledge of schistosomiasis was limited, as over one-third (35.35%) of participants did not know the cause of the disease. Misconceptions such as excessive salt intake and sun exposure were also common. Clinically, more than half of the participants reported hematuria (55.48%) and painful urination (51.23%), which are classical symptoms of UgS.

### Prevalence and intensity of *Schistosoma haematobium* infection

The overall prevalence of infection among the study participants was 34.23% (153/447), with a mean egg count (MEC) of 25.65 ± 1.74 eggs/10 mL of urine (Table 2). Most infected individuals had light-intensity infections (82.35%), while 17.65% had heavy-intensity infections.

**Table 2:**
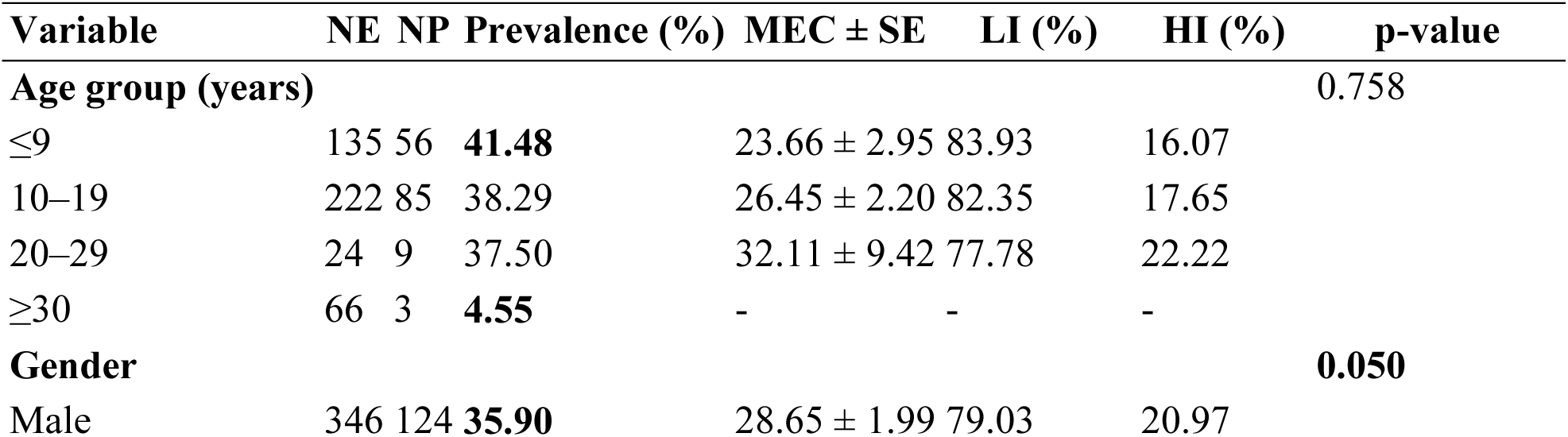
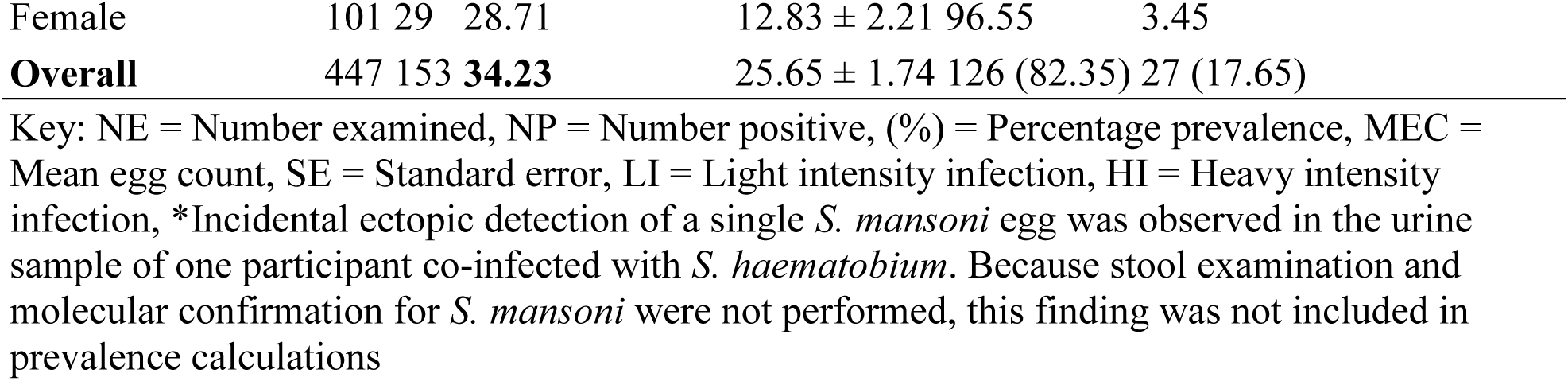
Prevalence and intensity of *Schistosoma haematobium* infection by demographic factors.

Across age groups, the highest prevalence was observed among participants aged ≤9 years (41.48%), followed by those aged 10-19 years (38.29%) and 20–29 years (37.50%). Participants aged ≥30 years had the lowest prevalence (4.55%). However, the association between age group and infection prevalence was not statistically significant (p = 0.76). Mean egg count was highest among participants aged 20-29 years (32.11 ± 9.42 eggs/10 mL urine), although most infections across all age categories were classified as light intensity.

With respect to gender, males recorded a higher prevalence of infection (35.90%) and higher mean egg count (28.65 ± 1.99 eggs/10 mL urine) compared with females, who had a prevalence of 28.71% and mean egg count of 12.83 ± 2.21 eggs/10 mL urine. Heavy-intensity infections were also more common among males (20.97%) than females (3.45%). The association between gender and infection prevalence was marginally significant (p = 0.05), suggesting that males were more exposed to infection risk factors than females.

The Prevalence of *S. haematobium* infection varied slightly among the communities, ranging from 32.35% in Maimanda to 37.84% in Danmadanhu. Danmadanhu recorded the highest prevalence (37.84%), followed by Gamji (35.90%) and Daddauda (34.12%), whereas Badau and Bangare each had a prevalence of 32.39% (Table 3).

**Table 3:** Community distribution of *Schistosoma haematobium* infection.

| Community | NE | NP | Prevalence (%) | MEC ± SE | p-value |
| --- | --- | --- | --- | --- | --- |
| Badau | 71 | 23 | 32.39 | $19.91 \pm 2.57$ | |
| Bangare | 71 | 23 | 32.39 | $33.43 \pm 4.26$ | |
| Daddauda | 85 | 29 | 34.12 | $23.83 \pm 3.25$ | |
| Danmadanhu | 74 | 28 | 37.84 | $14.82 \pm 3.46$ | |
| Gamji | 78 | 28 | 35.90 | $31.71 \pm 5.43$ | |
| Maimanda | 68 | 22 | 32.35 | $32.00 \pm 4.69$ | |
| **Overall** | **447** | **153** | **34.23** | **$25.65 \pm 1.74$** | **0.044** |

Key: NE = Number examined, NP = Number positive, (%) = Percentage prevalence, MEC = Mean egg count, SE = Standard error

Regarding infection intensity, Bangare (33.43 ± 4.26) and Maimanda (32.00 ± 4.69) showed the highest mean egg counts, while Danmadanhu had the lowest MEC (14.82 ± 3.46) despite recording the highest prevalence. The observed variation in infection intensity among the communities was statistically significant (p = 0.044), indicating that the burden of infection differed significantly across the study communities.

### Molecular characterization of *Schistosoma haematobium* isolates

PCR amplification of the mitochondrial *cox1* gene produced the expected 543 bp band in positive samples (Figure 3). To confirm *Schistosoma* species identity, sequence data were obtained from two isolates, DDA58 (Accession no. PX600373.1) and DMD02 (Accession no. PX600374.1) collected from Daddauda and Danmadanhu, respectively for confirmation of species identity. BLAST analysis of these *cox1* sequences showed 100% identity to *S. haematobium* reference sequence. Single nucleotide polymorphism (SNP) analysis showed that both isolates shared the same nucleotide pattern as the *S. haematobium* reference sequence and differed from *S. bovis* and *S. mansoni*. These findings support their identification as pure *S. haematobium* (Table 4).

**Figure 3:**
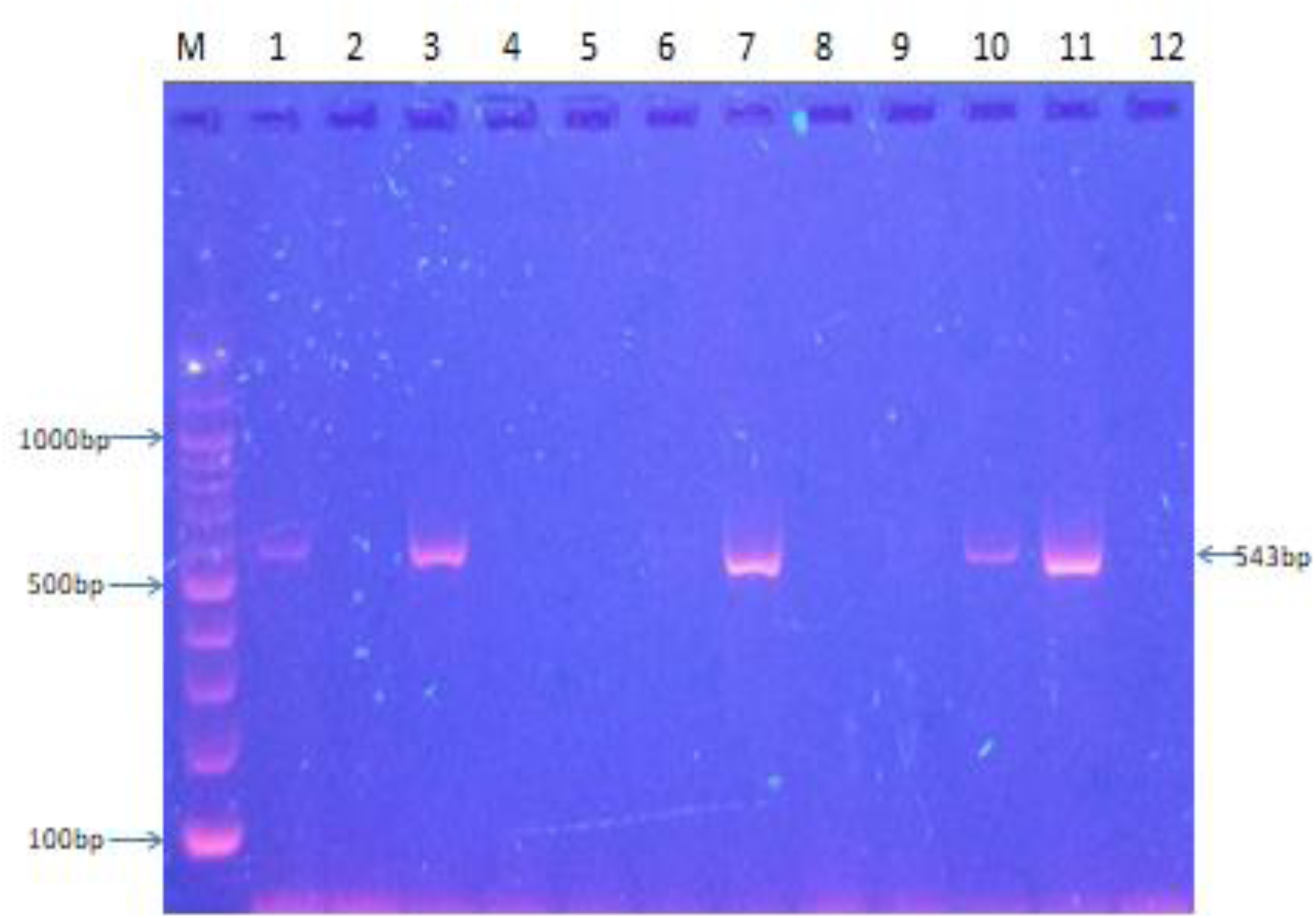
Molecular characterisation of *Schistosoma haematobium* mitochondrial *cox1* gene (543bp) in 2% agarose gel stained with 0.3 *µ*L ViSafe Gel Red. Lane M: standard size marker of 100bp. Bands in lanes 1, 3, 7 and 10 are positive samples; lane 11, positive control; lane 12, negative control

**Figure 4:**
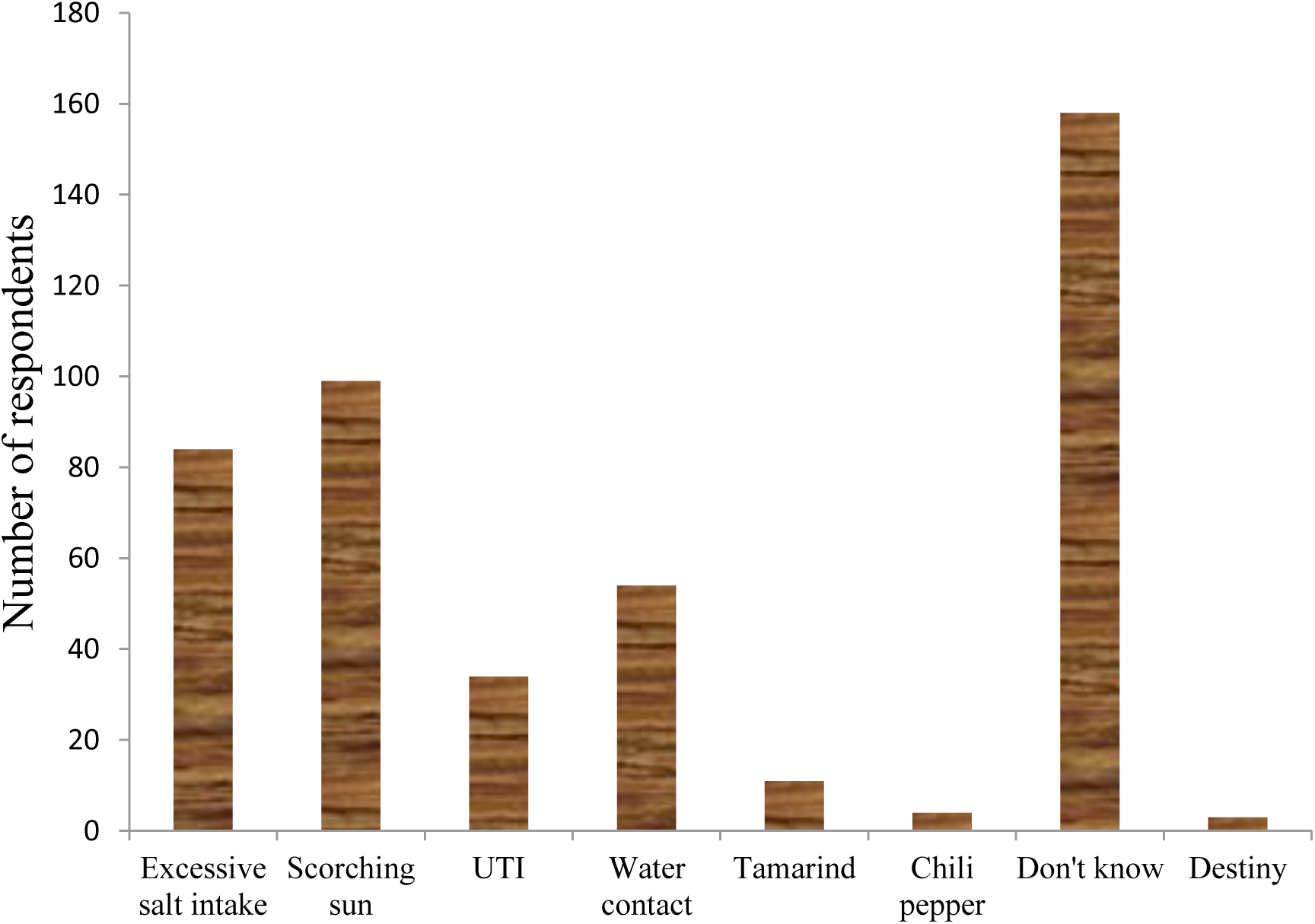
Perceptions on the etiology of *Schistosoma haematobium* infection among respondents in the study area

**Table 4:** Single nucleotide polymorphism (SNP) analysis of the mitochondrial *cox1* sequences from *S. haematobium* isolates.

| Accession number | *Schistosoma* spp. | SNP Positions |  |  |  |  |  |  |  |  |  |  |  |  |  |  |
| --- | --- | --- | --- | --- | --- | --- | --- | --- | --- | --- | --- | --- | --- | --- | --- | --- |
|  |  | 542 | 557 | 558 | 560 | 569 | 578 | 584 | 596 | 614 | 623 | 644 | 668 | 674 | 680 | 698 |
| MT579447.1<sup>*</sup> | *S. haematobium* | T | G | A | T | T | G | G | G | G | C | C | A | T | T | A |
| PX600373.1<sup>a</sup> | *S. haematobium* | T | G | A | T | T | G | G | G | G | C | C | A | T | T | A |
| PX600374.1<sup>b</sup> | *S. haematobium* | T | G | A | T | T | G | G | G | G | C | C | A | T | T | A |
| PP654276.1 | *S. bovis* | T | A | A | A | T | A | A | A | A | T | T | G | T | A | T |
| PQ177486.1 | *S. mansoni* | A | T | C | G | A | G | T | G | T | C | T | T | G | T | T |

\*Reference sequence, <sup>a</sup>DDA58 and <sup>b</sup>DMD02 isolates from the study area

### Sociodemographic and environmental determinants of urogenital schistosomiasis

The results in Table 5 present sociodemographic and environmental determinants of *S. haematobium* infection. Univariate analysis identified water contact as the strongest predictor of *S. haematobium* infection (OR = 18.96, 95% CI: 4.56–78.73, p < 0.001). Specific water-contact behaviors significantly associated with infection included swimming/playing (OR = 1.97, p = 0.003) and water fetching (OR = 1.85, p = 0.007).

**Table 5:**
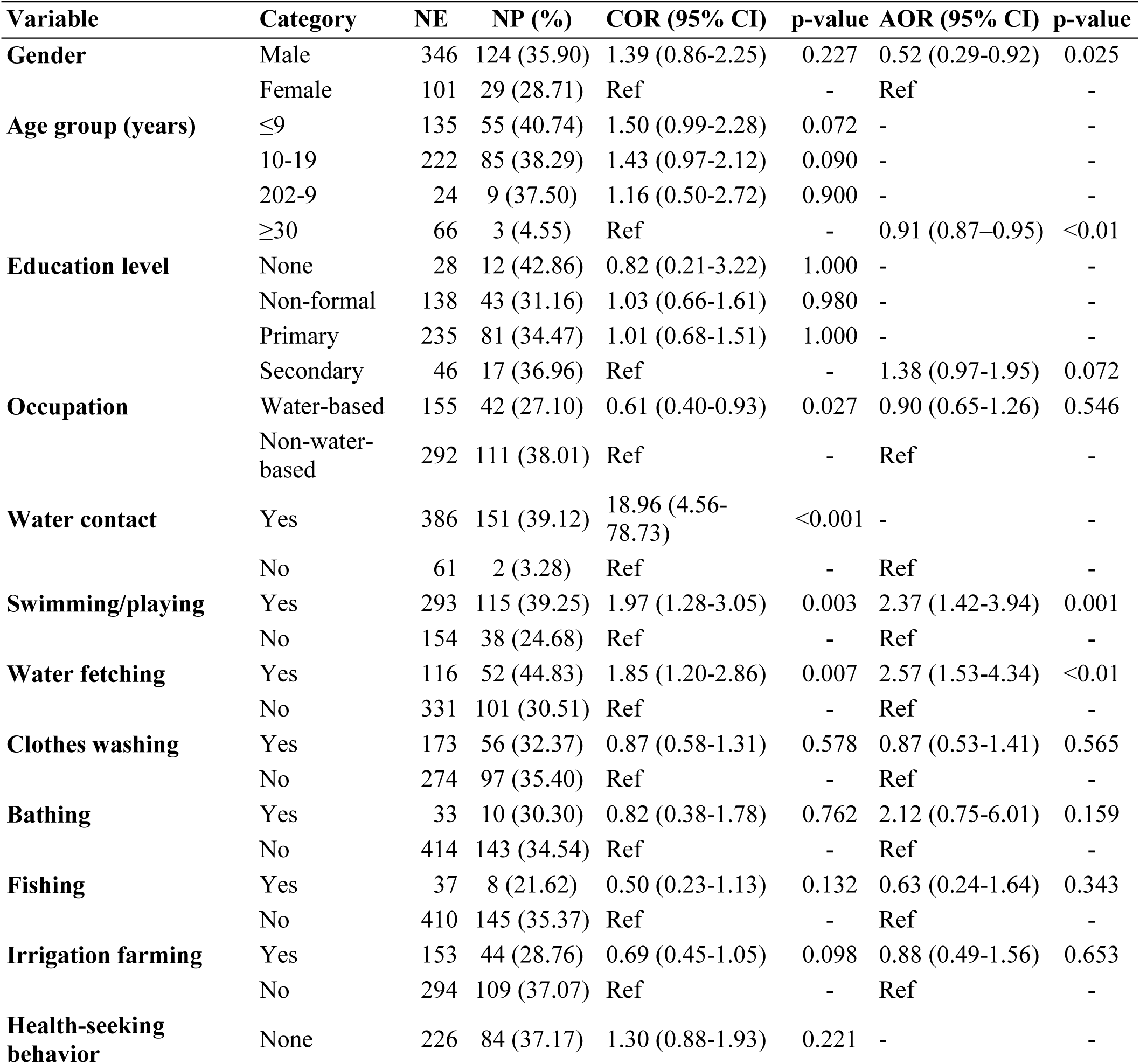
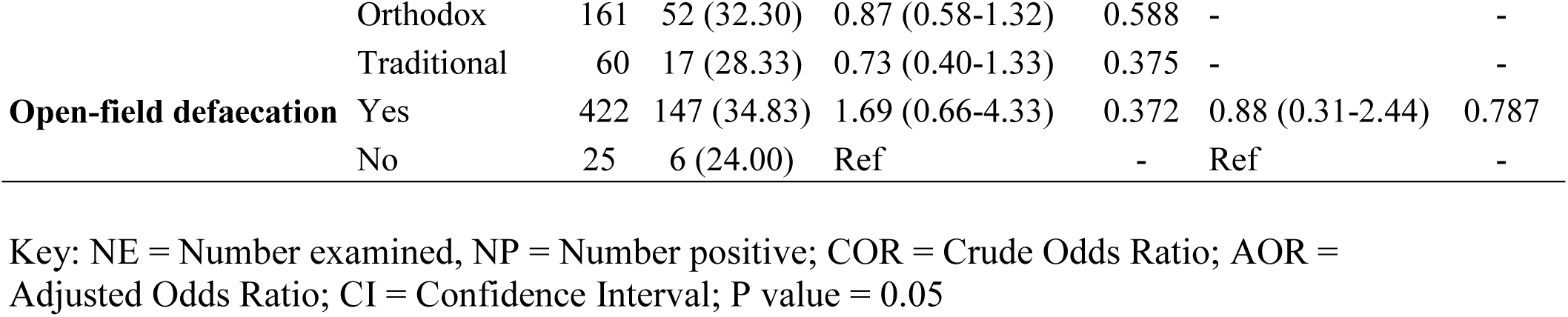
Univariate and multivariate logistic regression analyses of factors associated with *Schistosoma haematobium* infection among study participants (n = 447)

Participants involved in non-water-based occupations had significantly higher odds of infection than those engaged in water-based occupations (OR = 1.65, p = 0.027). Other variables, including gender, age group, education level, clothes washing, bathing, fishing, irrigation farming, health-seeking behaviour, and open-field defaecation, were not significantly associated with infection in the univariate model.

In the multivariate logistic regression analysis, gender (AOR = 0.52, 95% CI: 0.29–0.92; p = 0.025), age (AOR = 0.91, 95% CI: 0.87–0.95; p < 0.01), swimming/playing in water (AOR = 2.37, 95% CI: 1.42–3.94; p = 0.001), and water fetching (AOR = 2.57, 95% CI: 1.53–4.34; p < 0.01) remained significant predictors of infection. Participants involved in swimming/playing in water and water fetching had significantly higher odds of infection, while increasing age was associated with reduced odds of infection. Other variables were not independently associated with infection after adjustment for confounding factors (Figure 5).

**Figure 5:**
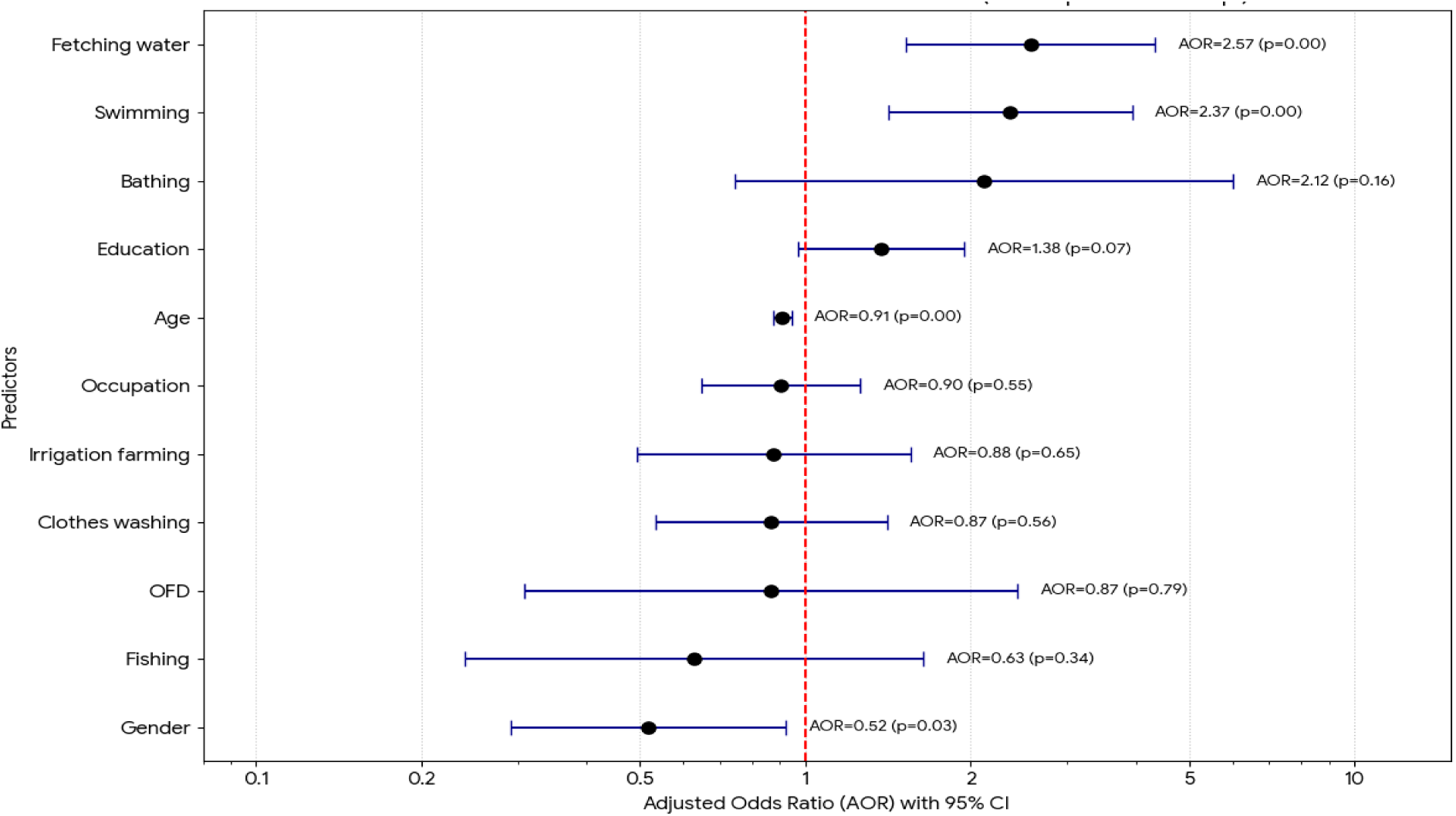
Forest plot of adjusted odds ratios for factors associated with *Schistosoma haematobium* infection among study participants

### Occurrence and infection status of *Bulinus* snail intermediate hosts

A total of 133 *Bulinus* snails were collected from the study area and examined for *Schistosoma haematobium* infection using cercarial shedding and PCR (Figure 6). *Bulinus truncatus* was the dominant species (117/133; 88.0%), while *Bulinus globosus* accounted for 16/133 (12.0%).

**Figure 6:**
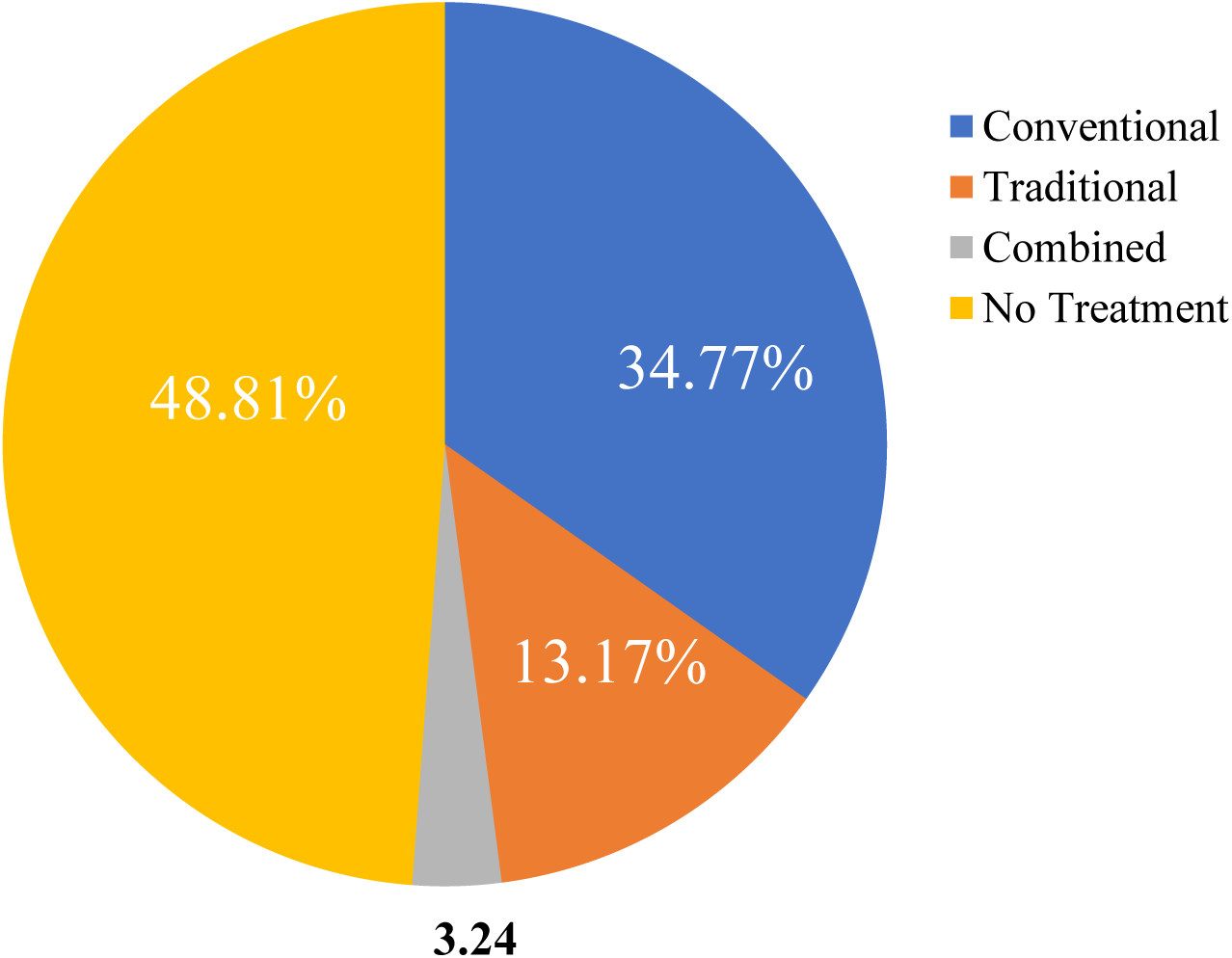
Schistosomiasis-related health-seeking behaviour of the study population

**Figure 7:**
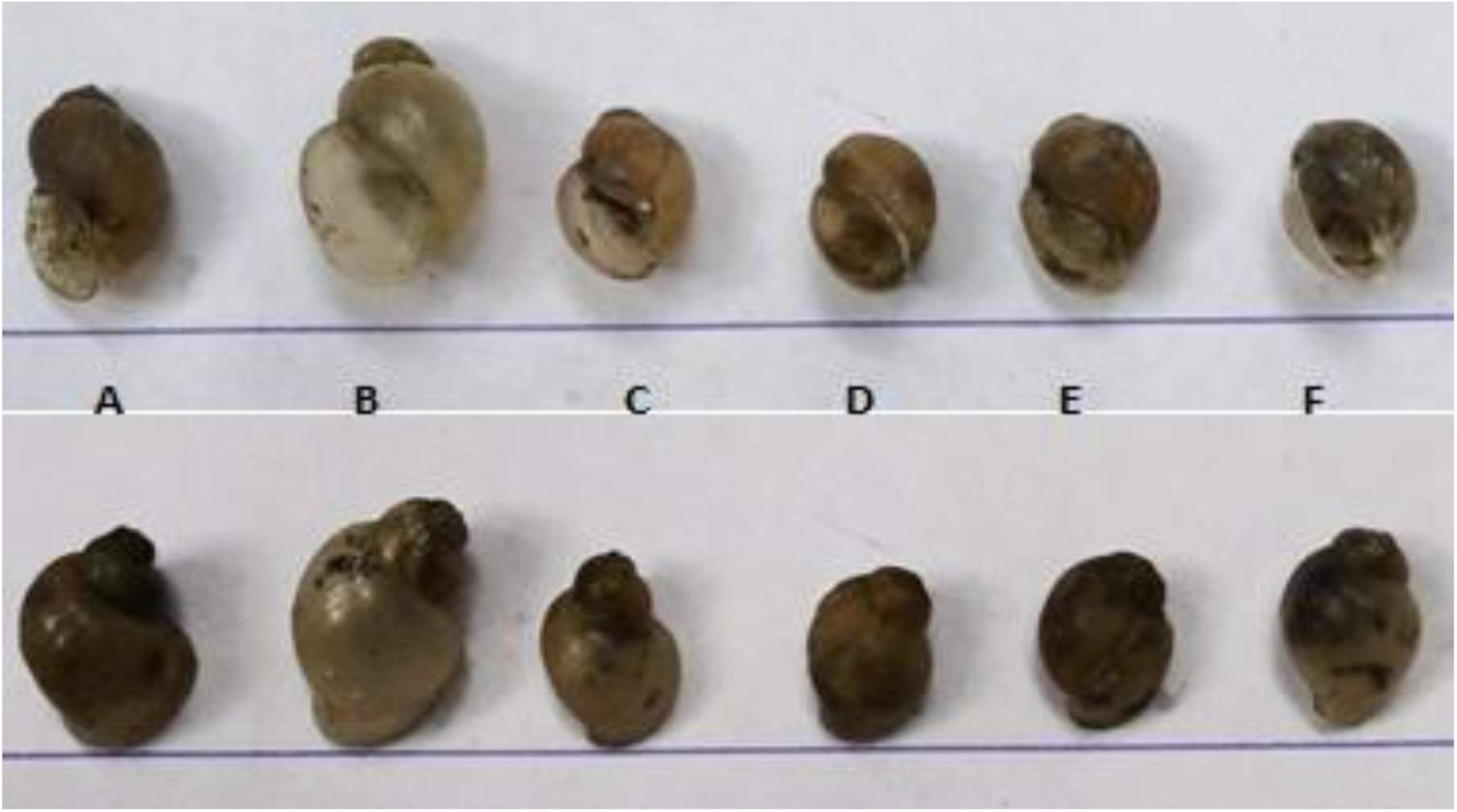
Apertural (upper row) and abapertural (lower row) views of *Bulinus* snails collected from the study area: A to C, *Bulinus truncatus*; D to F, *Bulinus globosus*

Overall infection prevalence was 3.8% (5/133) by cercarial shedding and 5.3% (7/133) by PCR, indicating higher sensitivity of PCR in detecting prepatent infections. Infection was unevenly distributed across sampling sites and species. In Bagwai, both species were infected. *Bulinus globosus* at Badau showed 50.0% prevalence (1/2), while *B. truncatus* at Daddauda had the highest infection rates (cercarial shedding: 14.3%; PCR: 21.4%). In Kafin Chiri and Wasai, infection occurred only in *B. truncatus* at Maimanda (4.6%) and Danmadanhu (cercarial shedding: 5.0%; PCR: 10.0%), respectively. There was a significant association between *Bulinus snail* infection status, species, and sampling site (χ² = 74.82; Fisher’s exact test P < 0.001) (Table 6). The small sample size for *B. globosus* at some sites should be interpreted with caution.

**Table 6:** Infection status of *Bulinus* snails with *Schistosoma haematobium* in the study area.

| Water reservoir | Sampling site | *Bulinus globosus* |  |  | *Bulinus truncatus* |  |  | Total |
| --- | --- | --- | --- | --- | --- | --- | --- | --- |
|  |  | N | NI1 (%) | NI2 (%) | N | NI1 (%) | NI2 (%) |  |
| Bagwai | Badau | 2(1.5) | 1(50.0) | 1(50.0) | 19(14.3) | 0(0.0) | 0(0.0) | 21 |
|  | Daddauda | 7(5.3) | 0(0.0) | 0(0.0) | 14(10.5) | 2(14.3) | 3(21.4) | 21 |
| Kafin Chiri | Gamji | 1(0.8) | 0(0.0) | 0(0.0) | 22(16.5) | 0(0.0) | 0(0.0) | 23 |
|  | Maimanda | 0(0.0) | 0(0.0) | 0(0.0) | 22(16.5) | 1(4.6) | 1(4.6) | 22 |
| Wasai | Bangare | 2(1.5) | 0(0.0) | 0(0.0) | 20(15.0) | 0(0.0) | 0(0.0) | 22 |
|  | Danmadanhu | 4(3.0) | 0(0.0) | 0(0.0) | 20(15.0) | 1(5.0) | 2(10.0) | 24 |
| Total |  | 16(12.0) | 1(6.3) | 1(6.3) | 117(88.0) | 4(3.4) | 6(5.1) | 133 |
| Overall prevalence (N=133) |  |  | NI1 = 3.8 |  |  | NI2 = 5.3 |  |  |

Key: N = Number of snails examined; NI1 = Number infected by cercarial shedding; NI2 = Number infected by PCR; (%) = Infection prevalence within species at each site; Statistics: $\chi^2 = 74.82$ ; Fisher's exact test $P < 0.001$ .

## Discussion

This study demonstrates sustained transmission of UgS in reservoir-adjacent communities in Kano State, Nigeria. The overall prevalence of *S. haematobium* infection (34.23%) indicates moderate endemicity according to WHO classification and reflects continued exposure to cercariae-infested water through domestic, occupational, and recreational activities. The relatively consistent prevalence across the six study communities suggests widespread transmission associated with shared ecological and behavioural risk factors, particularly dependence on reservoir water and frequent human–water contact. These findings reinforce the continuing relevance of the WHO neglected tropical diseases roadmap, which prioritises schistosomiasis control and transmission interruption in endemic settings (WHO, 2020; WHO, 2022).

Communities located around reservoirs in the present study represent a classic example of the poverty–disease nexus that underpins many neglected tropical diseases. Dependence on untreated reservoir water for domestic, occupational, and recreational activities reflects limited access to safe and affordable alternative water sources. Consequently, residents - particularly children and adolescents - remain repeatedly exposed to cercariae-infested water during activities essential for daily survival and livelihood. These findings suggest that UgS transmission in the study area is not solely driven by individual behaviour, but rather by entrenched socio-environmental inequities and inadequate rural infrastructure. This observation aligns with the broader conceptual framework of schistosomiasis as a disease of poverty and marginalisation in sub-Saharan Africa.

The prevalence observed in this study is comparable to previous reports from Kano State and other endemic regions of northern Nigeria, including the 37.0% prevalence reported from Wasai Dam by Enabulele *et al*. (2021) and 32.8% reported by Ali and Ndams (2013). Similar moderate prevalence levels have also been documented in Kebbi and Kwara States (Umar *et al*., 2017).

However, prevalence estimates across Nigeria remain heterogeneous, with some studies reporting substantially higher prevalence exceeding 40%–70%, while others documented lower prevalence below 10% (Duwa *et al*., 2009; Dawaki *et al*., 2015). These variations likely reflect differences in ecological conditions, water-contact behaviour, sanitation, treatment access, and population structure.

Although only two isolates were sequenced, molecular analysis confirmed the presence of non-hybrid *S. haematobium* based on mitochondrial *cox1* gene analysis. This provides preliminary evidence supporting species identity and absence of detectable hybridisation among the analysed isolates. Nevertheless, larger genomic studies involving multilocus sequencing are required to better characterise parasite population structure and possible introgression events in the region.

A rare incidental detection of *S. mansoni* eggs in urine was observed in one participant infected with *S. haematobium*. This may reflect ectopic egg elimination, mixed infection, or contamination during sample collection. However, because stool examination and species-specific molecular confirmation were not performed, definitive conclusions regarding co-endemicity or hybridisation cannot be established. Similar findings have previously been reported in Kano State (Enabulele *et al*., 2021).

Most infections in the present study were of light intensity despite the moderate prevalence observed. Similar predominance of light-intensity infections has been reported in northern Nigeria, including Jigawa State where Balogun *et al*. (2022) documented predominantly light infections and Kano State where Duwa *et al*. (2018) reported exclusively light-intensity infections among school children. The predominance of light infections may reflect repeated low-dose exposure, partial acquired immunity, or undocumented prior praziquantel exposure within the communities. Nevertheless, even light-intensity infections contribute substantially to transmission and chronic morbidity, including bladder pathology and female genital schistosomiasis. In contrast, Senghor *et al*. (2014) reported substantially higher proportions of heavy-intensity infections among school children in Senegal.

Age was an important determinant of infection, with the highest prevalence occurring among school-age children and adolescents. This pattern is consistent with the established epidemiology of schistosomiasis and is largely attributable to increased water-contact behaviour in younger individuals, particularly swimming, fetching water, and irrigation-related activities (Chipeta *et al*., 2013; Umar *et al*., 2017). The lower prevalence observed among older participants may reflect reduced exposure and gradual acquisition of partial immunity following repeated infections. These findings highlight school-age children as a priority target group for intervention while demonstrating that transmission extends beyond this population.

Although males had higher infection prevalence overall, interpretation of sex differences should be made cautiously because of the imbalance in male and female participation. The observed pattern likely reflects gender-related differences in exposure rather than biological susceptibility, as males are more frequently involved in outdoor and high-risk water-contact activities such as swimming, farming, and fishing-related tasks (Duwa *et al*., 2009; Dawaki *et al*., 2015). Importantly, infection among females remains a significant public health concern because of the long-term reproductive consequences associated with untreated UgS.

One of the strongest findings of this study was the significant association between infection and water-contact activities. Swimming and water fetching remained independent predictors of infection following multivariate analysis, emphasising the central role of direct exposure to untreated surface water in sustaining transmission. These findings are consistent with previous studies identifying human–water contact as the principal behavioural driver of *S. haematobium* transmission (Senghor *et al*., 2014; Trienekens *et al*., 2022). They further suggest that improvements in water infrastructure alone may be insufficient without sustained behavioural interventions aimed at reducing risky recreational and domestic water exposure.

Persistent transmission despite ongoing praziquantel-based mass drug administration (MDA) may reflect several interacting structural and programmatic challenges within the study communities. First, limited access to safe water compels residents to rely continuously on reservoirs for livelihood and domestic activities, thereby promoting rapid reinfection following treatment. Second, MDA coverage may remain suboptimal because school-based delivery approaches may inadequately reach high-risk populations outside formal educational systems, including Almajiri children enrolled in non-formal education settings. Third, poor access to recreational infrastructure encourages children to engage in swimming and play activities within cercariae-infested water bodies. Collectively, these factors suggest that preventive chemotherapy alone is unlikely to achieve sustained interruption of transmission without simultaneous improvements in water infrastructure, social services, and inclusive community-based treatment strategies.

The study additionally revealed substantial gaps in knowledge regarding schistosomiasis transmission and causation. Many participants attributed infection to misconceptions such as excessive salt intake, heat exposure, or dietary factors rather than contaminated water contact. Similar misconceptions have previously been reported among Hausa-speaking communities in Kano State (Dawaki *et al*., 2016; Umar *et al*., 2021). Importantly, poor knowledge should not be interpreted simply as individual negligence, but rather as evidence of long-standing underinvestment in culturally appropriate health education and risk communication within endemic communities. Poor disease awareness may contribute to delayed healthcare-seeking behaviour, normalisation of haematuria, and persistence of risky exposure practices. These findings further suggest that behaviour-change interventions focusing exclusively on individual responsibility may achieve limited success unless accompanied by structural interventions addressing inadequate water supply, sanitation, and social infrastructure.

Consistent with this, many infected individuals reported no formal healthcare-seeking behaviour, while reliance on traditional remedies remained common. Similar observations have been reported previously in Kano State, where traditional herbal treatment was often preferred over orthodox treatment for schistosomiasis-related morbidity (Umar *et al*., 2021). These findings suggest that chemotherapy alone may be insufficient for sustainable control unless accompanied by culturally appropriate health education and community engagement strategies.

Poor sanitary practices observed in the study communities may further contribute to transmission persistence. Open urination or defaecation near water-contact sites facilitates contamination of freshwater bodies with schistosome eggs, thereby sustaining the parasite life cycle (Nwabueze, 2013). These findings reinforce the importance of integrating water, sanitation, and hygiene (WASH) interventions into schistosomiasis control programmes.

Malacological findings further confirmed ongoing transmission within the study area. Two intermediate host species, *Bulinus globosus* and *Bulinus truncatus*, were identified, with *B. truncatus* representing the dominant and epidemiologically important species. Detection of infected snails by both cercarial shedding and PCR confirms active and prepatent transmission within the reservoirs. Similar occurrence of *Bulinus* snails has previously been reported from Wasai Dam and Kadawa irrigation areas in Kano State (Ali and Ndams, 2012; Duwa, 2017). The focal distribution of infected snails further suggests the existence of localised transmission hotspots contributing disproportionately to human infection.

The findings additionally highlight broader public health concerns associated with water-resource development projects in endemic regions. Artificial reservoirs may create ecologically favourable habitats for intermediate host snails while simultaneously increasing human dependence on unsafe water sources for irrigation farming, domestic use, and recreation. These interactions emphasise the importance of incorporating schistosomiasis surveillance and risk assessment into water-resource development planning.

The present findings additionally reinforce the relevance of a One Health framework for schistosomiasis control in reservoir-dependent communities. Reservoirs constructed to support agriculture and livelihoods may inadvertently create ecologically favourable habitats for intermediate host snails while simultaneously intensifying interactions among humans, livestock, and freshwater ecosystems. Although molecular analysis in the present study identified only non-hybrid *S. haematobium* isolates, the close ecological interface between humans, animals, and shared water bodies highlights the importance of integrated surveillance approaches incorporating human treatment, snail monitoring, environmental management, and veterinary considerations. Future studies involving larger sample sizes and multilocus genomic analyses may provide further insight into potential hybridisation dynamics within the region.

A major strength of this study is the integration of parasitological, molecular, behavioural, and malacological approaches within the same endemic communities, providing a more comprehensive understanding of transmission dynamics. However, several limitations should be acknowledged. The cross-sectional design prevented assessment of seasonal transmission dynamics, while use of a single urine sample may have underestimated prevalence, particularly among light infections. The underrepresentation of female participants may also have influenced sex-specific comparisons, and self-reported behavioural data may be subject to recall or social desirability bias.

## Conclusions

This study demonstrates sustained transmission of UgS in reservoir-dependent communities of Kano State, Nigeria, with moderate endemicity driven primarily by intense human–water contact, poverty-related reliance on unsafe water, and ongoing *Bulinus* snail infection. Molecular analysis confirmed the presence of non-hybrid *S. haematobium* among the analysed isolates. The integration of epidemiological, molecular, behavioural, and malacological evidence underscores the need for integrated control strategies combining preventive chemotherapy, WASH interventions, snail surveillance, health education, and community-based interventions to reduce reinfection and interrupt transmission within a broader One Health framework. These findings provide important baseline data for intervention planning and future ecological and molecular investigations in endemic communities.

## Author Contributions

Study conceptualization and design: MUA, SAL, ISN, EK,SK and JK. Field data collection and supervision: MUA, SAL, ISN, EK and JK. Laboratory investigation: MUA, ISN, GDC, SAL and SA. Data analysis and interpretation: MUA, SAL, ISN, EK, GDC, EOB and SA. Manuscript drafting, review and editing: MUA, ISN, GDC, EOB and SA.

## Data Availability

All data produced in the present work are contained in the manuscript

## Acknowledgements

The authors acknowledge the African Center for Neglected Tropical Diseases and Forensic Biotechnology, Ahmadu Bello University, and University of Bremen for financial and technical support through the Erasmus Scholarship of the European Union. Immense support from Aliko Dangote University of Science and Technology, Wudil and the Tertiary Education Trust Fund toward this doctoral fellowship is gratefully acknowledged. We thank Umar Aliyu Umar, Bello Adamu Jido, Muhammad Sabo and Bilya Auwalu Umar for their assistance in field data collection. The authors also appreciate Dr. Yu Choo Yee, Ms. Noof Bani Khaled and Ms.

Roshariza Haris from Universiti Putra Malaysia, for their technical support, as well as Dr. Ibrahim Mohamoudou for laboratory assistance at the Centre for Biomolecular Interactions Bremen, University of Bremen, and Natural History Museum, London, United Kingdom for their contributions. We especially appreciate Prof. Bonnie Webster for offering voluntary consultation during PCR troubleshooting and optimisation.

